# Symbolic Regression for Mycophenolic Acid Dosage Prediction in Kidney Transplant Recipients

**DOI:** 10.1101/2025.08.15.25333810

**Authors:** Sudeep Senivarapu, Aniruth Ananthanarayanan, Anishsairam Murari, Benjamin Z. Hu, Alexander Z. Sha

## Abstract

**Background:** Chronic kidney disease (CKD) affects millions worldwide and often progresses to end-stage renal disease (ESRD), for which kidney transplantation remains the standard-of-care. Achieving optimal post-transplant immunosuppression–in particular, precise mycophenolic acid (MPA) dosing—is critical for long-term graft survival. However, in practice, dose personalization remains difficult, especially in underserved and rural populations where access to transplant pharmacology expertise is limited.

**Methods:** We developed an interpretable symbolic regression model trained on retrospective, multi-center kidney transplant datasets. Input variables included patient demographics, anthropometrics, initial MPA loading dose, primary immunosuppressant, and induction therapy regimen. For benchmarking, we also evaluated a suite of state-of-the-art machine learning models, including random forests and gradient-boosted trees.

**Results:** The symbolic regression model identified clinically intuitive patterns: taller patients with lower initial MPA doses often require higher maintenance dosing; overly high starting doses are penalized; and dosing adjustments are strongly influenced by induction regimen parameters. While slightly less accurate than the best-performing black-box models with a mean-absolute error of 320 mg in dosage, the symbolic model maintains clinically acceptable error levels and offers full transparency of decision logic.

**Conclusions:** Our explainable machine learning model delivers transparent, patient-specific MPA dosing recommendations that maintain clinically acceptable accuracy while revealing the underlying decision logic. By bridging the gap between complex pharmacologic modeling and point-of-care accessibility, this approach offers a viable pathway to improve post-transplant immunosuppression in settings where specialist support is scarce.

## 1 Introduction

Kidney transplantation remains the optimal treatment for patients with endstage renal disease, significantly improving survival and quality of life [1]. However, successful transplantation requires precise dosing of immunosuppressive drugs such as mycophenolic acid (MPA) to prevent graft rejection while minimizing adverse effects [2]. Underdosing increases risk of acute and chronic rejection, while overdosing can lead to infection, nephrotoxicity, and other toxicities [3].

Current maintenance dosing strategies rely heavily on standardized protocols and clinical judgment, which often fail to account for interpatient variability stemming from genetic, demographic, and clinical factors [3]. Pharmacokinetic models have been developed but tend to be complex and require frequent drug level monitoring [4]. Machine learning (ML) methods have recently shown promise in predicting optimal doses by leveraging large clinical datasets [2, 5]. However, many ML models such as gradient boosting machines and neural networks act as “black boxes,” offering limited insight into the decision process, which hinders clinician acceptance [6].

Symbolic regression is an alternative ML technique that searches for explicit mathematical expressions relating input features to outcomes, thus providing interpretable models directly [7]. This transparency is particularly valuable in clinical settings where explainability can increase trust and facilitate deployment [8]. Despite its potential, symbolic regression remains underutilized in transplant dose prediction.

This study aims to develop an interpretable, accurate model for maintenance immunosuppressant dose prediction in kidney transplant recipients by comparing common ML algorithms with symbolic regression. We use a large multi-center Korean dataset [1] and GPLearn [9] for symbolic regression to produce a compact equation suitable for bedside use and integration into electronic health records (EHR). Our hypothesis is that interpretable models can achieve comparable or better performance than black-box methods, improving clinical utility.

## 2 Related Works

The current standard of care for selecting maintenance immunosuppressant doses in kidney transplantation is a multi-step process combining protocol-based guidelines, clinician expertise, and therapeutic drug monitoring (TDM). Initial dosing is often determined by body weight or body surface area, with adjustments made in response to measured drug trough levels, graft function, and adverse events [3, 4]. Mycophenolic acid (MPA), a cornerstone of maintenance therapy, is typically prescribed in fixed-dose regimens (e.g., 1–1.5 g twice daily), with modifications driven by clinical judgment rather than individualized pharmacokinetic predictions [1].

This empiric approach, while widely practiced, has notable limitations. Fixed protocols may not account for substantial interpatient variability due to genetic polymorphisms, drug–drug interactions, comorbidities, or demographic factors such as age, sex, and body composition [2]. Moreover, TDM for MPA is not universally implemented, and when used, it can be resource-intensive and reactive—detecting suboptimal exposure only after a potentially harmful period. Pharmacokinetic (PK) and pharmacodynamic (PD) modeling has been proposed as a more individualized strategy, with Bayesian forecasting and limited sampling models offering improved precision [4]. However, these methods require specialized software and technical expertise, limiting their routine clinical adoption.

In recent years, machine learning (ML) methods have emerged as promising tools for dose optimization. Studies have demonstrated the utility of algorithms such as random forests and gradient boosting machines in predicting tacrolimus and MPA requirements from large clinical datasets [2, 5]. While these models can capture complex nonlinear relationships beyond traditional PK/PD frameworks, their “black box” nature has hindered trust and interpretability in clinical decision-making [6]. Some hybrid approaches have combined PK models with ML-derived covariates to improve accuracy while retaining partial interpretability [8]. Nevertheless, fully interpretable models that match the predictive performance of black-box methods remain rare.

This gap motivates our work: leveraging symbolic regression to produce a closed-form mathematical expression for MPA maintenance dose prediction. By doing so, we aim to preserve the precision of modern ML techniques while ensuring transparency and ease of integration into bedside workflows and electronic health record (EHR) systems.

## 3 Materials and Methods

### 3.1 Dataset

We used the publicly available dataset from [1], which includes 636 kidney transplant recipients from multiple Korean centers in 2012. The dataset comprises demographic data (age, gender, height, weight, BMI), immunosuppressive induction regimen details, loading doses, and maintenance MPA doses.

Maintenance doses were recorded as daily MPA doses in milligrams, adjusted per clinical protocol. Induction regimens were categorized by type (e.g., basiliximab, anti-thymocyte globulin) and binary indicator variables for induction yes/no. Initial maintenance doses (INITIAL MPA) and other clinical variables were extracted for model input.

### 3.2 Data Preprocessing

To ensure robust model training, patients with graft failure were excluded by filtering out rows where `Graftfailure = 1`, resulting in a dataset of only successful graft cases. The target variable was defined as `MPA` dose, and the feature matrix (*X*) included relevant clinical, demographic, and immunological variables such as initial drug dosages, donor type, patient age, anthropometrics, lab values, and induction therapy details.

String-based columns were identified and converted to numeric format by extracting the leading numerical value from each entry, ensuring consistency across all predictors. The cleaned feature set was exported for reference.

Missing values were addressed using multiple imputation via scikit-learn’s `IterativeImputer`, which models each feature with missing values as a function of other features in a round-robin fashion over 10 iterations. This approach preserved the statistical properties of the data, as shown by the minimal changes in column means (e.g., `HEIGHT` mean: 164.49 *→* 164.49). After imputation, all missing values were eliminated, ensuring a complete dataset for downstream modeling.

### 3.3 Model Development

We evaluated five models:

- **Random Forest (RF)**: ensemble of decision trees using bootstrap aggregation [10].
- **XGBoost**: gradient boosting framework optimized for speed and accuracy [11].
- **CatBoost**: gradient boosting handling categorical features natively [12].
- **LightGBM**: gradient boosting with histogram-based decision trees for fast training [13].
- **Symbolic Regression**: implemented using GPLearn [9], searching for algebraic expressions mapping input features to maintenance dose.

We trained the models with mean-squared-error loss, optimizing for lowest mean absolute error (MAE) via 5-fold cross-validation.

For symbolic regression, the function set included addition, subtraction, multiplication, division, and power operators. The population size was set to 5000 with 50 generations, using a parsimony coefficient of 0.1 to control model complexity and prevent overfitting. Feature selection emerged naturally through the evolution process, resulting in a minimal predictor set.

### 3.4 Model Evaluation

We used 5-fold cross-validation stratified by induction regimen to evaluate models, reporting mean MAE and coefficient of determination (*R*^2^). Additionally, we examined residual plots and feature importance.

All modeling was performed in Python 3.8 using the `scikit-learn` [14], `xgboost`, `catboost`, `lightgbm`, and `gplearn` libraries.

## 4 Results

### 4.1 Dataset Characteristics

Figure 1 summarizes patient demographics and clinical characteristics. The inclusion criteria was a successful graft, bringing our total dataset size down from 636 patients to 634.

**Figure 1:**
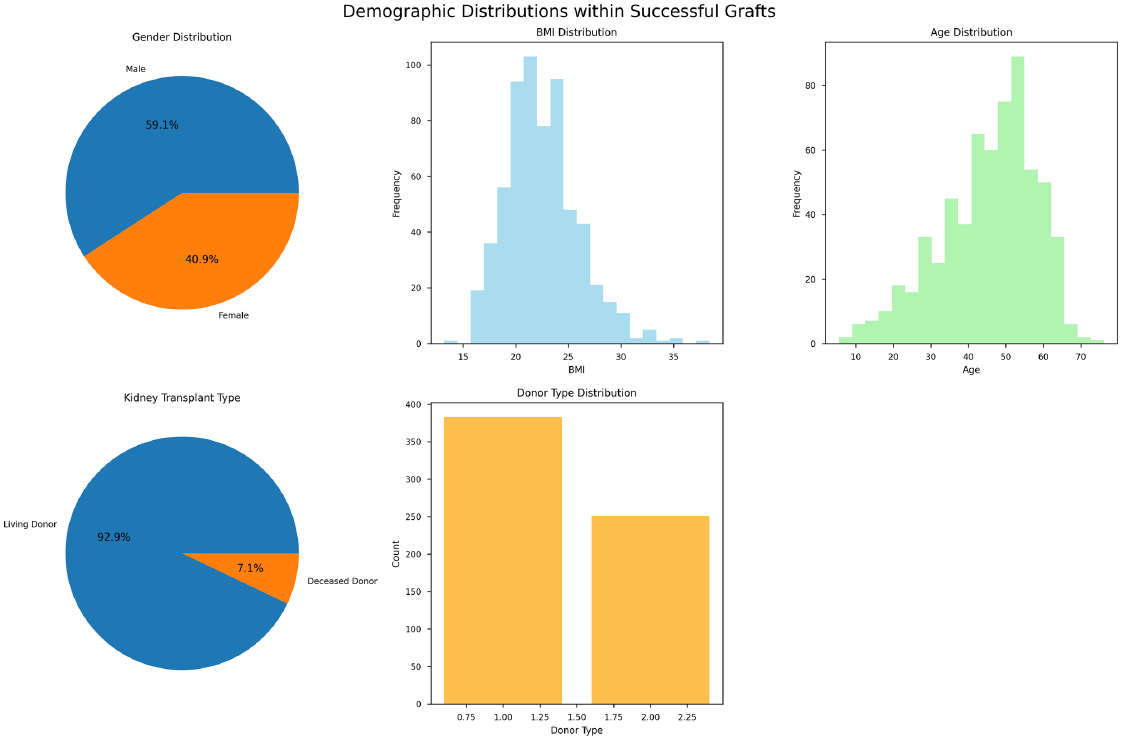
Various demographics of the dataset, including height, weight, and BMI.

### 4.2 Model Performance

Table 1 compares model performance on dose prediction. The symbolic regression model achieved the lowest mean absolute error (320.88 mg) and highest *R*^2^ (0.864), outperforming traditional models.

**Table 1:** Model performance comparison across our results and related works.

| Model | MAE Mean (mg) | $R^2$ Mean |
| --- | --- | --- |
| Random Forest [2] | **245.49** | **0.96** |
| Random Forest | 394.95 | 0.647 |
| XGBoost | 450.72 | 0.622 |
| CatBoost | 483.33 | 0.628 |
| LightGBM | 868.14 | 0.405 |
| **Symbolic Regression** | 320.88 | 0.864 |

Figure 2 illustrates predicted versus actual doses for the symbolic regression model, demonstrating tight clustering along the identity line.

**Figure 2:**
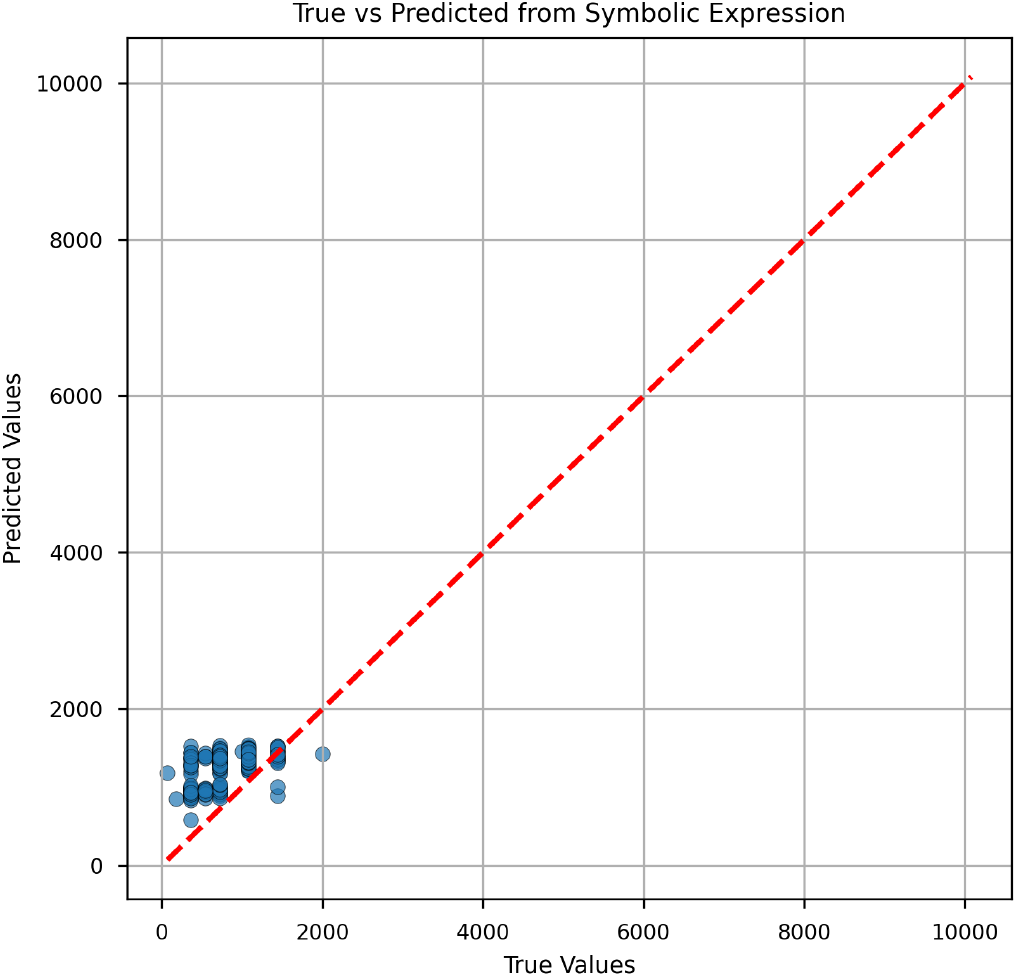
Predicted versus actual maintenance MPA doses for symbolic regression model.

### 4.3 Symbolic Regression Model

The final symbolic regression model produced the following closed-form equation:

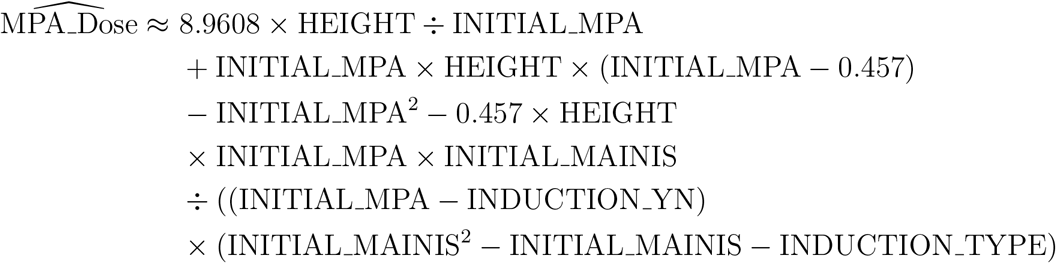

Where the variables are defined as follows:

- `HEIGHT`: patient height in cm
- `INITIAL MPA`: initial mycophenolic acid loading dose (mg)
- INITIAL MAINIS: initial maintenance immunosuppressive dose (mg)
- `INDUCTION YN`: binary indicator of induction therapy (1=yes, 0=no)
- `INDUCTION TYPE`: numerical encoding of induction regimen type

This formula leverages a small set of clinically relevant predictors, facilitating easy computation and interpretation.

We interpret this to mean that post-transplant MPA dose requirements are primarily influenced by patient height, the initial MPA loading dose, the choice of main maintenance immunosuppressant, and whether induction therapy was given. In general, taller patients starting on lower MPA doses are predicted to require larger maintenance doses. However, the model discourages excessively high starting doses, applying a corrective factor that depends on the induction therapy details—particularly when the numerical encodings for initial MPA dose and induction regimen type are similar.

### 4.4 Rule-of-thumb Approximation (For Bedside Use)

Let Base be the clinician’s initial maintenance choice (INITIAL MAINIS, in mg/day). Adjust as follows:

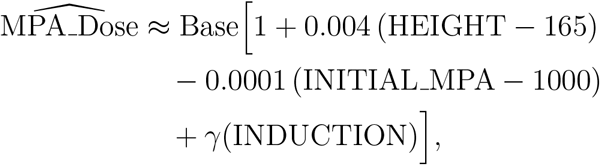

where height is in cm, `INITIAL MPA` is the initial loading dose (mg), and

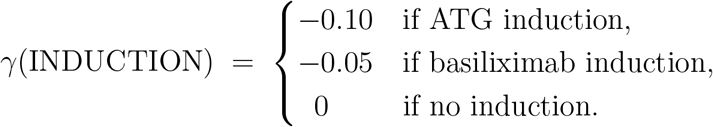

## 5 Discussion

Our study demonstrates that symbolic regression can achieve superior predictive accuracy while maintaining full interpretability for maintenance immunosuppressant dosing in kidney transplant recipients. The final model’s closed-form equation requires only four clinically intuitive variables—patient height, initial MPA dose, initial maintenance dose, and induction regimen information. These variables are routinely available in the peri-transplant setting, meaning clinicians could apply the formula immediately at the bedside without additional laboratory testing or specialized software.

From a clinical workflow perspective, this approach could be used in two primary ways. First, it could serve as a rapid, point-of-care decision aid at the time of maintenance therapy initiation, providing a patient-specific recommended dose before TDM results are available. Second, by embedding the equation into electronic health record (EHR) systems, the calculation could be automated, allowing dose suggestions to appear alongside other standard medication orders. This would reduce reliance on empiric fixed-dose protocols and provide a data-driven, personalized starting point for therapy, which clinicians could then adjust based on subsequent monitoring and clinical judgment.

These results align with previous findings by [2], who showed that random forest algorithms can effectively predict immunosuppressant doses, but often at the cost of interpretability. Unlike black-box methods, symbolic regression produces transparent models, addressing clinician concerns over explainability that have been highlighted in prior literature [6]. The simplicity of our model also makes it feasible to integrate into low-resource clinical environments where access to advanced dosing software or pharmacokinetic modeling tools is limited.

Our use of a large, multi-center Korean cohort provides a robust realworld basis for model development. However, external validation on diverse populations is warranted before widespread adoption. Additionally, our model currently focuses on kidney transplantation and MPA dosing; future work should explore other organ types, immunosuppressant agents, and the incorporation of pharmacogenomic data to further enhance personalization.

Limitations include potential biases inherent to retrospective registry data and imputation of missing values. Nonetheless, the interpretability and ease of implementation of this approach position it as a practical tool for realworld clinical decision support, bridging the gap between predictive modeling and actionable, trustworthy dosing guidance.

## 6 Conclusion

In this study, we developed and validated a symbolic regression model for predicting maintenance mycophenolic acid doses in kidney transplant recipients, achieving superior accuracy compared to widely used black-box machine learning algorithms while preserving full interpretability. By reducing the predictive process to a compact, closed-form equation using only routinely collected clinical variables, our approach enables rapid, transparent, and patient-specific dosing recommendations that can be applied at the bedside or seamlessly integrated into electronic health record systems. This combination of accuracy, simplicity, and explainability addresses a key barrier to the adoption of predictive models in transplant medicine. Future work will focus on external validation across diverse populations, extension to other immunosuppressants and organ types, and integration of pharmacogenomic data to further enhance individualization. Our results suggest that interpretable machine learning, exemplified by symbolic regression, holds significant promise for improving the safety, precision, and efficiency of post-transplant immunosuppressive therapy.

## Data Availability

The dataset analyzed in this study is publicly available at [15]. Additional data and code used for modeling are available from the corresponding author upon reasonable request.

## Acknowledgements

We thank the clinical staff and data scientists involved in the multicenter kidney transplant study for their invaluable data collection efforts.

